# Framework for Implementation of Poisson MaxSPRT Technique with Variations for Vaccine Safety

**DOI:** 10.1101/2024.07.24.24310962

**Authors:** Md Samiullah, Jim Buttery, Hazel J. Clothier, Jiying Yin, John Mallard, Jeremiah Munakabayo, Gonzalo Sepulveda Kattan, Gerardo Luis Dimaguila

**Affiliations:** Murdoch Children’s Research Institute; Royal Children’s Hospital, 50 Flemington Rd, Parkville, VIC - 3052, Australia; Australian Financial Security Authority, Australia

**Keywords:** Continuous Surveillance, Adverse events, Vaccine pharmacovigilance, Sequential Probability Ratio, Signal detection, reporting delay, real-world datasets

## Abstract

For direct, continuous, and sequential drug and vaccine safety surveillance, the Maximized Sequential Probability Ratio Test (MaxSPRT) was developed by the Centers for Disease Control and Prevention (CDC) (Kulldorff et al, 2011). Its predictive value and power to detect signals and the ability to monitor adverse events continuously have made it an emerging technique for vaccine adverse event surveillance. Moreover, being able to use a statistical method e.g. MaxSPRT in the absence of dose distributed denominator is a practical advantage for spontaneous reporting systems to function as stand-alone signal detection systems. In this paper, we present a comprehensive framework for implementing MaxSPRT for vaccine safety surveillance and Poisson data. We analysed the literature regarding MaxSPRT and sequential analysis. Our analysis revealed numerous variations of MaxSPRT, adapted to the specific requirements and objectives of the users. Variations are due to differing types of data and purpose of use, including whether used for epidemiological surveillance or for regulatory monitoring. This paper provides a comprehensive guide for organisations contemplating the implementation of MaxSPRT. It synthesises existing literature on MaxSPRT, identifies variations based on specific requirements, and describes an implementation framework. We offer a detailed explanation of the steps and challenges associated with the implementation of MaxSPRT on the adverse event following immunisation (AEFI) reporting database of Surveillance of Adverse Events Following Vaccination in the Community, Victoria, Australia (SAEFVIC), the largest jurisdictional reporting service by volume in Australia. It also proposes some techniques and measures to deal with the challenges associated with the implementation process.

**Key Points:** MaxSPRT is a powerful method for ongoing vaccine surveillance, offering flexibility to adapt to various situations and data limitations. However, this flexibility can lead to challenges in implementation. Our paper simplifies the MaxSPRT method with clear explanations and step-by-step guidance, addressing potential issues and proposing solutions to improve its use in monitoring vaccine and drug safety.

## 1. Introduction

Vaccines keep us safe against harmful diseases by increasing the natural defense of our body and inducing immunity to various infections (Murphy et al, 2023). Every licensed vaccine goes through rigorous testing over numerous trial phases before being approved for use by regulators. Nonetheless, adverse events following immunisation could occur, although they are typically modest and transient, like a painful arm or a low-grade fever. Though relatively rare, more severe side effects may occur. Following approval, vaccine safety surveillance services continue to scan and analyse data from a variety of sources for any signs that a vaccine might have unexpected harmful consequences on health, or an increase in incidence or severity of known reactions, known as post-licensure surveillance.

In order to perform post-licensure vaccine safety signal detection, researchers propose diverse approaches that involve various statistical methods, techniques, and measures in original, modified, or combined forms. Research and vaccine surveillance groups may also impose necessary modifications to the approaches during implementation, addressing their organisational strengths and limitations, availability of data, and resources.

Key statistical measures and techniques used in post-licensure vaccine surveillance include Proportional Reporting Ratio (PRR) (Clothier et al, 2019), Cumulative Sum (CUSUM) (Mesfin et al, 2021), Reporting Odds Ratio (ROR) (Yoon et al, 2020), Empirical Bayesian Geometric Mean (EBGM) (Lee et al, 2020), Chi-Square, and MaxSPRT (Kulldorff et al, 2011)-a specialised variation of Sequential Probability Ratio Test (SPRT) (Wald, 1992) proposed by Wald.

An important aspect of vaccine surveillance is continuous monitoring of adverse events that vaccinees may experience as vaccines are administered, through a continuous sequential analysis that detects a signal when adverse events exceed the probability that they occur by chance (Kulldorff et al, 2011). Sequential Probability Ratio Test (SPRT) (Wald, 1992) was proposed by Wald to detect adverse events through hypothesis testing. It is used to determine the most likely hypothesis among a set of hypotheses and is appropriate for sequentially independent and identically distributed (iid) data, which implies that the data comes from the same distribution. SPRT was then refined for vaccine surveillance, and we refer to this as Classical SPRT (cSPRT) in this paper. cSPRT is used for surveillance to test if the null hypothesis (no vaccine safety signal or the vaccine is safe), is more likely than the alternative hypothesis (there is a vaccine safety signal). However, cSPRT is highly sensitive to the relative risk chosen in the alternative hypothesis construction, which makes it difficult to use for ongoing, routine surveillance.

Consequently, Kulldorff et al. proposed the use of a maximised sequential probability ratio test (MaxSPRT) (Kulldorff et al, 2011) based on a composite alternative hypothesis, which works across a range of relative risks. However, the various, specific objectives and data sources of vaccine surveillance groups have added to the complexity of implementing them using the different measures and parameters required for MaxSPRT analysis. This subsequently resulted in various implementations of MaxSPRT by other researchers in the community.

To our knowledge, there is currently no literature describing the variations of MaxSPRT implementation, detailed and step-wise calculations, along with when to use them based on vaccine surveillance objectives, and data sources and types.

## 2. Methods

This section describes the evolution of SPRT and MaxSPRT as reported in the literature. It forms the foundation for the framework proposed in this paper for the implementation of MaxSPRT technique variations for vaccine safety.

In both Wald’s SPRT and cSPRT (Wald, 1992), and Kulldorff’s MaxSPRT (Kulldorff et al, 2011), the number of adverse events is considered to be random, while the cumulative person-time or the cumulative number of vaccinations, if the follow-up time per vaccination is uniform, is considered to be fixed. The predicted number of adverse events under the null hypothesis is then considered to be a known function of the total persontime and certain possible confounding variables like age, sex, and location.

Based on the dependence and sufficiency of the comparator data MaxSPRT can be divided into two groups, (i) Conditional MaxSPRT and (ii) unconditional MaxSPRT. When the background information is insufficient and conditional on an event, Kulldorff developed conditional MaxSPRT (cMaxSPRT) (Center for Biologics Evaluation and Research, 2021), where it factors in the number of adverse events in the historical data and the surveillance population and computes the cumulative person-time it took to observe so many events as the random variable.

The unconditional MaxSPRT can be further subdivided based on the properties of the dataset into two types (Kulldorff et al, 2011; Center for Biologics Evaluation and Research, 2021; Li and Kulldorff, 2010): (i) pMaxSPRT for Poisson distribution data where values are real numbers, comparator data are not conditional on any event and sufficient background information is available; and (ii) bMaxSPRT for data with a binomial distribution where the SPRT analysis is conducted for each event being observed that is binary for each event. The trend of variations continues for some further factors and parameters. These make the calculation of MaxSPRT quite challenging due to numerous variations. The parameters, hyperparameters, and conditions that need to be considered while calculating MaxSPRT are: (i) the grouping of the reports based on age; (ii) the decision to use any comparator rate, sources of comparator rate, the process of calculating the comparator rate; (iii) the length of the risk window and the decision to slice or not slice risk windows; (iv) the decision to use the length of exposure, written as “*l*_*stiwa*_” in the figure (Figure 1) and later denoted as *l*_*stiwa*_ in the rest of the paper, as a proxy to the calculation of exposed person time, and the variations in calculating the exposed person-time; (v) the choice of using a test margin in the calculation; and (vi) the decision to use a delay adjustment, as well as the ways to calculate the delay adjustment.

**Fig. 1.**
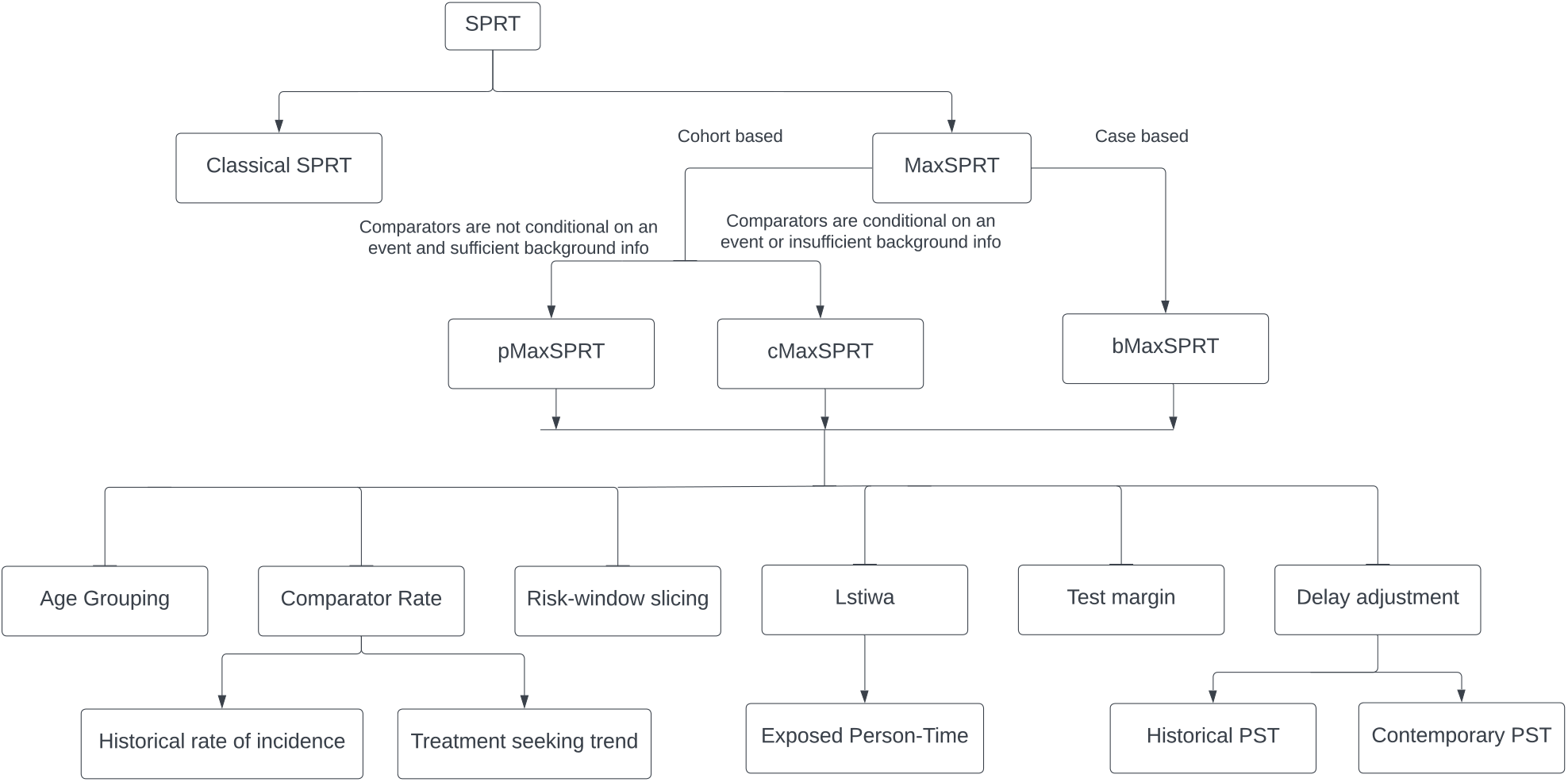
Different variations of MaxSPRT.

**Fig. 2.**
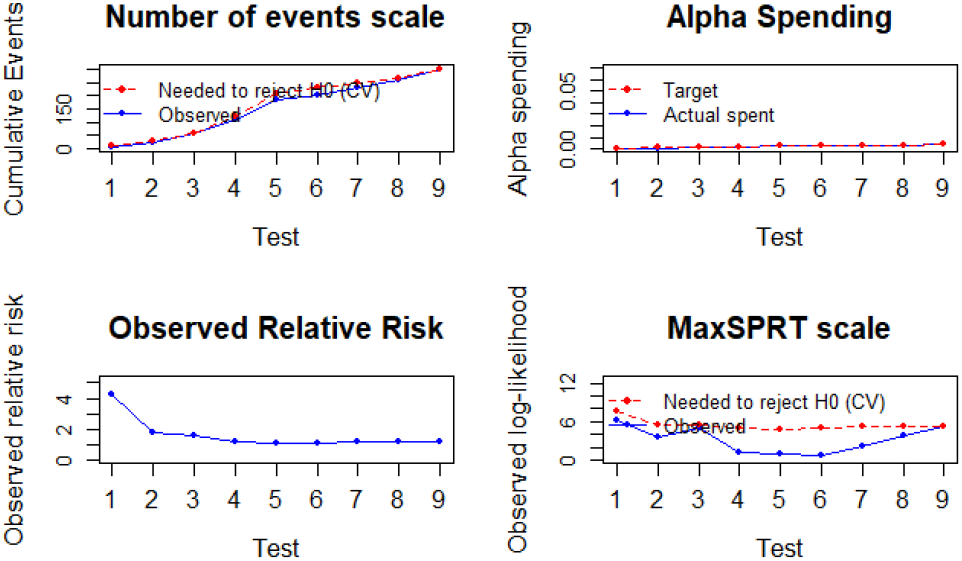
Visualising signal detection using MaxSPRT.

Each combination of the aforementioned factors makes a different variation of MaxSPRT, therefore, the factors and hyperparameters are introduced briefly as follows and described in greater detail with relevant technical information below.

### Age grouping

Sometimes, we can stratify the whole MaxSPRT analysis based on age groups. In such cases, all the calculations are performed at a more granular level and all the hyperparameters, such as the historical comparator rate, are taken based on age grouping. For example, age strata can be 5-year or 10-year age groups, or bespoke grouping according to vaccine schedule. More small-ranged grouping can also be performed to address the risk of different AESIs for different age ranges. Moreover, unlike the static binning detailed above, a dynamic binning approach of risk and AEFI-based age grouping is also possible where all groups may not have the same interval. This could provide an option of sequential analysis. This kind of analysis has not been tried yet in the literature, to the best of our knowledge.

### Comparator group

One of the hyperparameters, which is also known as the historical comparator group. This is required as a reference value in calculating the likelihood of an event. This value sometimes needs scaling down based on the length of the observed period. There are several ways of calculating this rate, and the information available at the time of analysis also plays an important role. In the case where more than one option is available, the most relevant and reasonable one needs to be used. Two such potential options, as per (Kulldorff et al, 2011), are:

- *Retrospective rate of incidence*: This approach calculates the rate of incidence of the same AEFI in previous years. This rate can be calculated from the historical data, reflecting the ratio of reports in the previous years.
- *Treatment-seeking trend*: In this approach, the rate is calculated from General Practitioners’ (GP) which is a term used in Australia that refers to primary care in general, clinics’, and hospitals’ data and health records by finding the trend of treatment-seeking behaviour for a particular AEFI.

### Risk-window slicing

In MaxSPRT, while calculating various metrics, we can take the elapsed time of an observation period into consideration. This helps mitigate the gap of overestimating the risk or control period of a vaccine. It also allows slicing the risk window into smaller periods so that an observation period that is shorter than the risk window can scale down the required metrics as per the length of the observation period. In the sliced version, we further stratify the data, especially reports for different intervals of the risk window. This provides a more granular level of calculation and tuning. In (Greene et al, 2011), authors show the efficacy of using a sliced risk window.

### *l*_*stiwa*_ calculation

the measure required to estimate the expected number of events if the dose distribution information is unknown or insufficient.

### Exposed person-time

The Centers for Disease Control and Prevention, USA (CDC) in (Center for Biologics Evaluation and Research, 2021) proposed a technique and a formula to calculate the length of exposure in a person-time unit by capturing the trend of overlapping exposed time for the vaccinees within the risk window.

### Other alternative estimations

If the CDC’s approach can be replaced by some more reasonable approach that can capture the length of exposure more realistically and effectively.

### Test margin

This is an optional hyperparameter that can be used to fine-tune the estimated expected number of events. For each AESI within each database, the test margins are defined based on comparator rates derived from historical data of the outcome, adjusted for the length of the risk window and a target number of doses required to cause harm. This specification aims to enhance the operating characteristics of the pMaxSPRT and improve the quality of information available for regulatory decision-making. To perform a one-sided test where the null hypothesis states that the rate of AESI in the vaccinated group is not higher than the historical comparator rate by a specific test margin (*m*), where *m* is a non-negative value stated as a percent of the comparator rate.

Hence, the choice of using it or not using it makes another fold of variations in MaxSPRT calculation, and of course, needs an assessment to determine which one is a better choice: using it or not using it.

Moreover, in the case of using it, finding the optimal value of the test margin is also a challenge.

### Delay adjustment

AESIs are not reported in real-time for various reasons. In sequential and continuous surveillance, this factor must be addressed in any version of MaxSPRT to adjust the observed number of events within the observation period. This adjustment to the observed number of events is required to help MaxSPRT find appropriate values for CVs to decide on the likelihood of signals in the study.

There are several ways of calculating and adjusting the delay especially estimating the delay from retrospective analysis. Based on the availability of the data or reports in previous years for the same AEFI and for the same vaccines, we have divided the ways into two high-level classes.

### Historical p(s, t)

If the analyser has access into the reports for the AEFI and the vaccine under consideration, then CDC’s approach (Center for Biologics Evaluation and Research, 2021) as explained in later sections (Section 3.2.3) can be followed to calculate *p*(*s, t*) for dealing with delay adjustment.

### Concurrent p(s, t)

Where the analyser does not have historical information on the trend of delays in reporting, such as in the case of a new AEFI or a new vaccine, the delay adjustment must be done by some predictive or approximation approach using the so far obtained concurrent data. We also explain this and propose a method to deal with this situation in Section 3.2.3.

## 3. Results

After analysing the relevant literature, we developed a taxonomy of variations of MaxSPRT as shown in Figure 1.

The structure and organisation of the taxonomy depict the factors that establish the SPRT subtypes and implementation variations in a hierarchical manner. The variation starts with the availability of baseline information required to do the computation for surveillance. In some systems and situations, the expected number of events comes with associated uncertainty. There are various reasons behind the uncertainty that can add additional challenges to surveillance. To deal with uncertainty in the projected expected number of events, a conditional maximised sequential probability ratio test (cMaxSPRT) (Li and Kulldorff, 2010) was proposed by Li et. al., with a known baseline risk function no longer needed. Instead, it considers the unpredictability that comes from both the surveillance population and historical data.

To aid readability and implementation, we list the terminologies used in the literature to construct the narrative of the paper clearly. Table 1 enlists the terms, their notations, and symbols that are commonly used throughout the paper. We refer to Appendix A for the definitions, examples, and explanations of the terms.

**Table 1.** Notations

| Terms | Notations |
| --- | --- |
| Study week | $s$ |
| Any week within the study period | $t$ |
| Exposure weeks | $w$ |
| Time to onset | $TTO$ |
| Slices in Risk window | $\delta$ |
| Expected number of events | $\mu_s$ |
| Observed number of events | $Y_s$ |
| Historical comparator rate | $\theta$ |
| Per day comparator incidence rate of age-group $a$ | $\theta_a$ |
| Proportion of data complete | $p(s, t)$ |
| Observed exposed person-time | $L_{stiwa}$ |
| Log-likelihood Ratio | $LLR$ |

### 3.1. Properties of the Dataset

SAEFVIC (a Surveillance of Adverse Events Following Vaccination in the Community data) (Clothier et al, 2017) is a set of timely passive surveillance data consisting of AEFI reports submitted by vaccinees, their relatives, GPs, nurses, or other healthcare professionals. It aligns to minimum data for AEFI surveillance as per WHO (Organization et al, 2014). Each row of the data contains a report converted to a transactional database format where each report is identified by a unique identifier named *V accinee ID*. Corresponding to each unique identifier, there are the age and sex information of the vaccinee, the time stamps of the dose administration, reporting time (when was it posted), and report submission. It also contains the time to onset expressed in the number of days for the adverse event followed by the description of the event, reaction sign/symptom or clinical term of the event name, and vaccine brand as well as the dose received. There are other data that are neither relevant nor required for MaxSPRT analysis, and we filtered them in the preprocessing step.

Like other real-world datasets, our data have missing values too. This needs special attention before the data can be used for analysis. Data are collected through an online interactive form that is used by endusers (reporters). Hence, it is prone to errors like human errors that include misspellings and typographical errors. This implies that we need to use some mechanism to deal with unclean data.

Table 2 shows the headers of a sample, a synthetic dataset developed from real-world data after altering it so that MaxSPRT analysis can detect a signal for demonstration purposes. We used this sample data for the analysis with selected columns, the corresponding data types, and the domains of the columns.

**Table 2.** SAEFVIC data set used in this paper with their attributes, types, and domains of the attributes, with pseudo sample data.

| Attributes | VacID | Vaccine Name | Age | Sex | Time of Vaccination | TTO | Reporting Time | Submission Time | Reactions |
| --- | --- | --- | --- | --- | --- | --- | --- | --- | --- |
| Types | String | String | Number | Binary | Time stamp | Number | Time Stamp | Time Stamp | String |
| Domains | Report ID | Name | Years | M/F | Time<br>Format:<br>dd/mm/yyyy HH:MM:SS | Days | Time<br>Format:<br>dd/mm/yyyy HH:MM:SS | Time<br>Format:<br>dd/mm/yyyy HH:MM:SS | Set of names<br>Comma Separated |
| Semantics | Unique Identifier | Name of the<br>vaccine brand | Age of the<br>Vaccinee at<br>vaccination | Gender<br>of the<br>Vaccinee | Time of dose<br>administration | Time to<br>On setting<br>the AEFI | When the AEFI<br>was reported | When the report was<br>recorded in SAEFVIC | The Adverse Event |

### 3.2. Steps and processes of implementation

Here we explain step by step the calculation of each measure, factor, and parameter required to calculate *LLR* (Log-likelihood Ratio) which is used to detect a signal in MaxSPRT analysis. Throughout the paper, as a running worked example to demonstrate the calculations, we have chosen a sample SAEFVIC dataset for a particular vaccine and “Abdominal pain” as an AEFI. Refer to Appendix B (Implementation) for full details of each step, particularly during a MaxSPRT implementation in the SAEFVIC setting.

#### 3.2.1. Computing Cumulative Observed Number of events

We have taken all the reports into account after preprocessing (refer to Appendix B.1, Preprocessing) the data. For each week 1 ≤ *t* ≤ *s*, and each age group *a*, we counted the number of reports, say *Y*_(*t,a*)_. Summing all the counts for all age groups for a particular week, *t*, gives the number of observed events for the week. We added a column *Y*_*a*_ for each age group *a*’s report count and two more columns to the data, one for keeping track of the weekly observed report count *Y*_*t*_ and the other one for keeping the cumulative counts of observed reports from week 1 up to a corresponding week *t, Y*_1,*t*_.

As an illustration, Table 3 shows the weekly count of observed events with a breakdown based on age strata. The last column shows the cumulative count of events.

**Table 3.** Demonstration of computing observed report count for MaxSPRT analysis for Abdominal Pain

| Computing Observed Report count for MaxSPRT analysis for Abdominal Pain |  |  |  |  |  |  |  |  |  |  |  |
| --- | --- | --- | --- | --- | --- | --- | --- | --- | --- | --- | --- |
| #Week | Report count<br>of age 0-9 | Report count<br>of age 10-19 | Report count<br>of age 20-29 | Report count<br>of age 30-39 | Report count<br>of age 40-49 | Report count<br>of age 50-59 | Report count<br>of age 60-69 | Report count<br>of age 70-79 | Report count<br>of age 80+ | Weekly<br>Total<br>Report | Cumulative<br>Report<br>Count, $Y_t$ |
| 1 | 0 | 0 | 2 | 2 | 3 | 2 | 0 | 0 | 0 | 9 | 9 |
| 2 | 0 | 0 | 4 | 2 | 3 | 3 | 2 | 1 | 0 | 15 | 24 |
| 3 | 0 | 0 | 7 | 12 | 7 | 3 | 2 | 1 | 1 | 33 | 57 |
| 4 | 0 | 0 | 8 | 13 | 11 | 8 | 5 | 5 | 1 | 51 | 168 |
| 5 | 0 | 0 | 17 | 16 | 18 | 14 | 7 | 5 | 1 | 78 | 246 |
| 6 | 0 | 0 | 0 | 1 | 0 | 4 | 6 | 6 | 2 | 19 | 265 |
| 7 | 0 | 0 | 0 | 2 | 1 | 7 | 6 | 7 | 1 | 24 | 289 |
| 8 | 0 | 0 | 1 | 1 | 1 | 10 | 14 | 3 | 1 | 31 | 320 |
| 9 | 0 | 0 | 0 | 1 | 0 | 20 | 18 | 2 | 0 | 41 | 361 |
| 10 | 0 | 0 | 0 | 0 | 1 | 4 | 1 | 2 | 0 | 8 | 369 |

**Table 4.** Demonstration of computing *l*_*stiwa*_ for MaxSPRT analysis for Abdominal Pain

| Computing $l_{stiwa}$ for MaxSPRT analysis for Abdominal Pain | | | | | | | | | | |
| --- | --- | --- | --- | --- | --- | --- | --- | --- | --- | --- |
| #Week | $l_{stiwa}$<br>(0-9) | $l_{stiwa}$<br>(10-19) | $l_{stiwa}$<br>(20-29) | $l_{stiwa}$<br>(30-39) | $l_{stiwa}$<br>(40-49) | $l_{stiwa}$<br>(50-59) | $l_{stiwa}$<br>(60-69) | $l_{stiwa}$<br>(70-79) | $l_{stiwa}$<br>(80+) | Weekly<br>$l_{stiwa}$ |
| 1 | 0 | 0 | 2 | 2 | 5 | 1 | 0 | 0 | 0 | 10 |
| 2 | 0 | 0 | 18 | 5 | 13 | 9 | 2 | 0 | 0 | 47 |
| 3 | 0 | 0 | 24 | 18 | 37 | 10 | 8 | 4 | 3 | 104 |
| 4 | 0 | 0 | 51 | 57 | 51 | 38 | 14 | 28 | 15 | 254 |
| 5 | 0 | 0 | 76 | 79 | 84 | 52 | 23 | 25 | 7 | 346 |
| 6 | 0 | 0 | 10 | 12 | 22 | 7 | 17 | 26 | 2 | 96 |
| 7 | 0 | 0 | 0 | 3 | 6 | 11 | 30 | 14 | 1 | 65 |
| 8 | 0 | 0 | 6 | 7 | 0 | 33 | 24 | 16 | 1 | 87 |
| 9 | 0 | 0 | 7 | 8 | 1 | 91 | 53 | 12 | 0 | 172 |
| 10 | 0 | 0 | 7 | 7 | 4 | 41 | 13 | 10 | 0 | 82 |

#### 3.2.2. Length of exposure calculation

The length of exposure is used to estimate the expected number of events in some variations of MaxSPRT analyses. This is roughly the cumulative count of exposed time per vaccinee after a dose is administered and until the reaction is set on them in the person-time unit. Definition 7 (Appendix A, Terminology) expresses a way of capturing the overall length of exposure for all vaccinee under consideration for a MaxSPRT analysis when dose administration information is not available at all, or at least not during the analysis in real-time.

#### 3.2.3. Proportion of data complete calculation

AESIs are not reported in real-time for various reasons. Even after being reported, AEFI recording may also be delayed. This phenomenon is known as observation delay. Due to such delays, at the time of analysis, the amount of the observed data for a particular study week is likely to be less than the actual amount. This leads to an overestimation of the expected number of events, which imposes a potential risk of missing a signal. To address the issue, MaxSPRT has a feature of approximation and this is known as calculating the proportion of data complete (the ratio of the observed and expected amount of data). We defined the term in Definition 14.

As we discussed in Appendix A, CDC defined *p*(*s, t*) in (Center for Biologics Evaluation and Research, 2021). However, the process of calculation or estimation of the term *p*(*i*) was not clearly stated, and neither was it illustrated clearly in the paper. Although it looks quite straightforward to calculate this value from the ratios of reports obtained and expected, when we tried to apply the method we ended up with several possible ways of calculating the value. This adds more factors and hence variations of MaxSPRT calculation. We also propose a new but similar way of calculating the proportion of data complete. We describe an equation to estimate the proportion of data complete that can be used to adjust the factor of delays in reporting incidents in Section 3.2.7 using concurrent data.

#### 3.2.4. Computing comparator rate, θ

There are several ways of calculating this rate and two of the plausible ways are described in Appendix A. In our current implementation, we calculated this value retrospectively from the previous years’ data (Andrews et al, 2023). We have utilised our available data from previous years of the same reaction for calculating the background rates and used the formula mentioned in Definition 11 and Definition 12. These background rates were extracted and analysed by our epidemiology team.

#### 3.2.5. Test Margin, M

To the best of our knowledge, this is a value first used by CDC (Center for Biologics Evaluation and Research, 2021) for MaxSPRT analysis. It is particularly used to fine-tune the estimation of the expected number of events *µ* and used to emphasise a particular AEFI and vaccine/drug pair. Although, in our implementation, we facilitate specifying this value, by default we have used 1.0 for *M*. In particular, this can be a value close to 1 such as 1.01.

#### 3.2.6. Estimating the Expected number of events

The expected number of events is the number of events expected to be observed without rejecting the null hypothesis. Calculating this number is not straightforward. It depends on the available information. There is a significant difference in calculating or estimating this value based on the available information. An approach to estimate this measure is proposed in (Center for Biologics Evaluation and Research, 2021) and defined in Definition 8 as “Approximate Expected number of events” in Appendix A.

In passive surveillance or reposting systems, such as SAEFVIC, we do not have dose administration information in real time. In such systems, only the numerator, and the report count are available. Therefore, in this paper and in our current implementation, we focused on *l*_*stiwa*_ (observed exposed person-time, see Appendix A, Terminology) based estimation of the denominator. Once all the required metrics for calculating the expected number of events (*µ*_*s*_) are found, we compute the expected count using Definition 8 (Appendix A, Terminology). While processing the data, another column is added to it to keep the weekly cumulative expected number of event counts that is going to be utilised later to compute *LLR*.

#### 3.2.7 Calculation of Data Complete

We propose a new but similar way of calculating the proportion of data complete. We describe an equation to estimate the proportion of data complete that can be used to adjust the factor of delays in reporting incidents. We specified the way of calculating the ratio. The denominator of the ratio calculation is subject to the choice of dataset and timeline we choose.

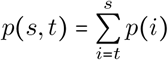

where, 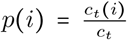, and *c*_*t*_ is the total number of reports for week *t, c*_*t*_(*i*) is the total number of reports for week *t* at week *i*.

In several variations of MaxSPRT, using this factor is optional, such as in (Greene et al, 2011). In case we decide to use this factor, the calculation of the proportion of observed data can be performed in two ways. Firstly, similar to the existing works (Center for Biologics Evaluation and Research, 2021; Greene et al, 2011), we can use historical data for the same AEFI and the same vaccine or similar vaccine. Secondly, where no historical data is available, a special approach should be adopted to calculate the *p*(*i*) using the concurrent data by estimating the proportion of later weeks of exposure from the earlier weeks of exposure.

In order to estimate the proportion of data complete from concurrent data, we can use the trend of delays in reporting for the weeks up to the study week. As an illustration of the reporting delay distribution, see Table 5. This distribution is for a MaxSPRT analysis on week 10 observed in an analysis performed to check for a safety signal for Abdominal pain. Note that the count of reports for each week with corresponding report submission weeks. The table contains the distribution of AEFI reporting with certain delays. As an example, it is expected that there will be 6 reports for week 3 and those were reported in weeks 3, 5, 6, and 7. Similarly, 25 reports of weeks 4, and 5 were reported in week 4, 10 on week 5, 5 on week 6, 2 on week 7, and 1 on each of week 9, 10, and after 10 weeks.

Refer to Appendix D to check the detailed calculation of the ratio to get *p*(*s, t*). This metric is used in the MaxSPRT analysis and demo calculations.

#### 3.2.8 Calculating LLR and detecting signals

We used cumulative expected number of events, *µ*_*s*_ (a snapshot of calculation of all components/measures of *µ* is shown in Table 6) and *Y*_*s*_ up to week *s*, to calculate the *LLR* using the formula in Definition 17. Either, the *LLR* or *Y*_*s*_ is used to detect signals by comparing against the critical value, *CV*. Note, there are two types of critical values: (i) based on *LLR* of the corresponding week’s cumulative expected number of events, we denote it as 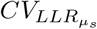 and (ii) based on *Y*, the corresponding week’s observed event count, we denote it as 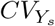. In other words, say in week *t*, if 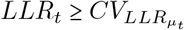 or 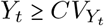, then there is a signal.

**Table 5.** Demonstration of Reporting delay distribution & computing *p*(*s, t*) for Abdominal Pain

| Reporting delay distribution & computing $p(s, t)$ for Abdominal Pain | | | | | | | | | | | |
| --- | --- | --- | --- | --- | --- | --- | --- | --- | --- | --- | --- |
| AEFI Occuring Week | Reported on Week 3 | Reported on Week 4 | Reported on Week 5 | Reported on Week 6 | Reported on Week 7 | Reported on Week 8 | Reported on Week 9 | Reported on Week 10 | Reported after Week 10 | Total Reports | Total Reports up to week s [s=10] |
| 3 | 1 | 0 | 2 | 2 | 1 | 0 | 0 | 0 | 0 | 6 | 6 |
| 4 |  | 5 | 10 | 5 | 2 | 0 | 1 | 1 | 1 | 25 | 24 |
| 5 |  |  | 11 | 33 | 10 | 2 | 2 | 0 | 10 | 68 | 58 |
| 6 |  |  |  | 23 | 27 | 9 | 1 | 2 | 10 | 72 | 62 |
| 7 |  |  |  |  | 31 | 38 | 9 | 0 | 10 | 88 | 78 |
| 8 |  |  |  |  |  | 7 | 10 | 2 | 2 | 21 | 19 |
| 9 |  |  |  |  |  |  | 3 | 6 | 16 | 25 | 9 |
| 10 |  |  |  |  |  |  |  | 9 | 18 | 27 | 9 |

**Table 6.**
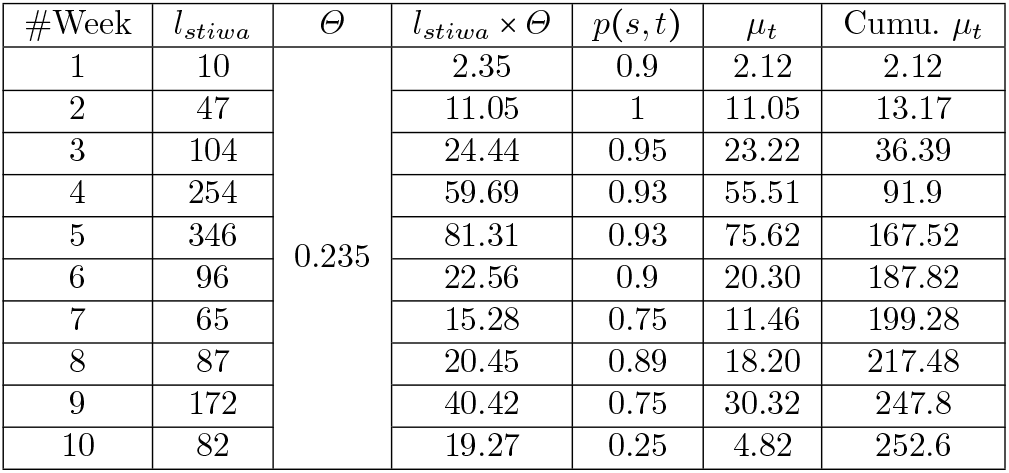
Computing required parameter values for MaxSPRT analysis for Abdominal Pain using the formula in Definition 8

| #Week | $l_{stiwa}$ | $\Theta$ | $l_{stiwa} \times \Theta$ | $p(s, t)$ | $\mu_t$ | Cumu. $\mu_t$ |
| --- | --- | --- | --- | --- | --- | --- |
| 1 | 10 | 0.235 | 2.35 | 0.9 | 2.12 | 2.12 |
| 2 | 47 |  | 11.05 | 1 | 11.05 | 13.17 |
| 3 | 104 |  | 24.44 | 0.95 | 23.22 | 36.39 |
| 4 | 254 |  | 59.69 | 0.93 | 55.51 | 91.9 |
| 5 | 346 |  | 81.31 | 0.93 | 75.62 | 167.52 |
| 6 | 96 |  | 22.56 | 0.9 | 20.30 | 187.82 |
| 7 | 65 |  | 15.28 | 0.75 | 11.46 | 199.28 |
| 8 | 87 |  | 20.45 | 0.89 | 18.20 | 217.48 |
| 9 | 172 |  | 40.42 | 0.75 | 30.32 | 247.8 |
| 10 | 82 |  | 19.27 | 0.25 | 4.82 | 252.6 |

At this stage, let’s demonstrate the detection of the presence of any signal in a dataset. As shown in Table 7, the cumulative *Y*_*t*_ and *LLR*_*t*_ for study week, *t* = 9 of the data is, respectively, 301 and 5.34 where 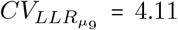 for *µ*_9_ = 247.8 (obtained from (Kulldorff et al, 2011, Table 1) and not shown in Table 7) and 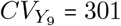 (obtained from Poisson MaxSPRT analysis using R Sequential package (Silva and Kulldorff, 2024) with a sample size = 500, total *α* = 0.05 with 10 installments for 10 tests, each test having *α* = 0.005, *ρ* = 0.5, the minimum number of events = 1). Hence, MaxSPRT indicates a signal in the week, *t* = 9, with a 98% likelihood that the vaccine is causing abdominal pain.

**Table 7.** Demonstration of signal detection through MaxSPRT analysis for Abdominal Pain (null hypothesis, *H*_0_ = no signal and when *H*_0_ is rejected, there is a signal)

| Week#, $t$ | Test# | $\mu_t$ | $Y_t$ | Cumu. $\mu_t$ | Cumu. $Y_t$ | $RR_t$ | $LLR_t$ | $CV$ | Reject $H_0$? |
| --- | --- | --- | --- | --- | --- | --- | --- | --- | --- |
| 1 | 1 | 2.12 | 9 | 2.12 | 9 | 4.25 | 6.13 | 10 | No |
| 2 | 2 | 11.05 | 15 | 13.17 | 24 | 1.82 | 3.57 | 27 | No |
| 3 | 3 | 23.22 | 33 | 36.39 | 57 | 1.57 | 4.97 | 58 | No |
| 4 | 4 | 55.51 | 51 | 91.9 | 108 | 1.18 | 1.33 | 124 | No |
| 5 | 5 | 75.62 | 78 | 167.52 | 186 | 1.11 | 0.98 | 209 | No |
| 6 | 6 | 20.3 | 19 | 187.82 | 205 | 1.09 | 0.76 | 233 | No |
| 7 | 7 | 11.46 | 24 | 199.28 | 229 | 1.15 | 2.11 | 247 | No |
| 8 | 8 | 18.2 | 31 | 217.48 | 260 | 1.2 | 3.91 | 267 | No |
| 9 | 9 | 30.32 | 41 | 247.8 | 301 | 1.21 | 5.34 | 301 | Yes |
| 10 | 10 | 4.82 | 8 | 252.6 | 309 | 1.22 | NA | NA | NA |

Note in the above example, to perform MaxSPRT, especially using the Sequential package of R language, we need to provide some values as parameters: Sample size, Statistical significance (*α*) and alpha spending plan, alpha spending function (*ρ*), statistical power, the minimum number of events, the observed number of events, and the expected number of events. Calculation of the expected number of events is highlighted in the paper. To aid the readers, we briefly introduced those terms in Appendix E.

### 3.3. Challenges and Lessons Learnt

MaxSPRT was introduced to conduct continuous sequential analysis that detects potential signals when adverse events exceed the probability that they occur by chance, and allows for continuous monitoring of adverse events that may occur in temporal association following immunisation. However, Continuous surveillance for signal detection through MaxSPRT is challenging. The measure itself is quite complex and has been used and implemented in various ways. The challenges can be divided into three stages: 1. pre-implementation, 2. implementation, and 3. post-implementation.

Before we started the implementation of MaxSPRT, we faced a number of challenges. The first challenge was establishing a clear understanding of the procedure, terms, and underlying mathematics due to its many variations. The variations depend on the availability, nature, and properties of the data in hand. In addition, there is no hard and fast rule or specified model for the implementation. Organisations have different capabilities and limitations that direct implementers towards different variations, as explained in Section 1.

The challenges we faced during implementation were mainly due to the lack of information in the literature. Most papers described the analysis process from a high level, with limited detailed information about the implementation. Although some papers such as (Center for Biologics Evaluation and Research, 2021; Mesfin et al, 2019) explain in more detail, they are not fully reproducible. As an example, the definition of *p*(*s, t*) is insufficiently defined to reliably reproduce (as discussed in Section 3.2.3). The data source and technique to calculate the “proportion of data complete” that captures and addresses the observation delay in reporting reactions add several variations in MaxSPRT, and dealing with the calculation of this term was challenging.

Calculating the expected number of events was the most onerous. Its significance in the whole surveillance process increases the difficulty because any significant mistake or error in the calculation of this term will mislead the surveillance. It has several components such as *p*(*s, t*), *l*_*stiwa*_, *θ* and each adds different variations (discussed in Section 1 and Section 3.4), especially when we do not have any prior information about the admissibility and correctness of the approaches and heuristics we have adopted.

Deciding whether to use age stratification or not, and if used, choosing between static grouping or dynamic binning technique adds to the challenge. Moreover, in the case of static grouping, deciding which grouping would give us optimal and more indicative results is not known and is subject to empirical analysis. In the case of either type of grouping, calculating the background rates for each age group also adds another layer of difficulty.

Interestingly, the data source and technique to calculate the background rates add some variations and thus additional challenges are introduced. Data properties and characteristics such as the distribution of the data also increase complexity in the implementation. One of the significant inherent challenges in any process involving data is missing values. In the case of MaxSPRT analysis, missing information in the data needs special consideration. We handled the missing data with great care, explained in Appendix B.1.

We also had to deal with some challenges regarding the selection of the development and implementation tool and setting up the environment. We explained the issue in detail with our workaround in Appendix B.3.

Moreover, it is also challenging to validate or assess if the implementation of MaxSPRT was correctly performed; to the best of our knowledge, there is no common and publicly available data to be used as a benchmark and standard outcome to compare the results due to the privacy, security, and ethics policies of the data used in the literature. Choosing and using the right version of MaxSPRT adds to the complexity. To deal with all of these challenges, we implemented the framework in a way that allows the end user to exhibit different variations of MaxSPRT for analysis using some arguments, and adjustment or changes to their values. We listed the available variations in Section 3.4.

### 3.4. Incorporating variations

MaxSPRT exhibits numerous variations that we explained in Section 1. In this section, we explain what variations are allowed in our current implementation. From the beginning, our focus was to make the implementation as flexible as possible so that the end user could utilise different variations of MaxSPRT by tuning the values of the relevant parameters.

The main goal of this research was to provide support for vaccine safety analysis even in case sufficient baseline information is not handy. For this purpose, we follow the calculation of the expected number of events proposed by CDC (Center for Biologics Evaluation and Research, 2021). Their calculation of the expected number of events is termed in this paper as Approximate Expected Number of Events as defined in Definition 8

(Appendix A, Terminology). The equation in the definition takes into account several parameters, such as *l*_*stiwa*_, *p*(*s, t*), *α*, and *M*. Each of these parameters introduces different variations of MaxSPRT, as discussed in previous sections (such as Sections 1 and Appendix B), at least because of different approaches available and adapted by various research groups in their implementations of the parameters and MaxSPRT. Moreover, if we follow the approach of (Greene et al, 2011), we can add more variations. From our point of view, the following variations can be applied, and we provide the flexibility to use all the variations in our current implementation.

1. 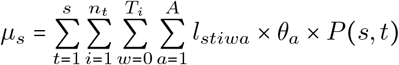
2. 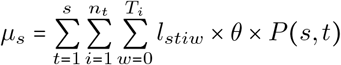
3. 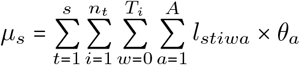
4. 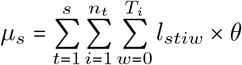

Note that in Option 1, we have not used the test margin unlike CDC’s original calculation (Center for Biologics Evaluation and Research, 2021). We can also vary the calculation by not using any age stratification for *θ* and thus for other parameters, as we did in Option 2 and Option 4. We also omitted the use of *p*(*s, t*) in Option 3 and Option 4. More such variations could be introduced by taking the combination of all four parameters used in Definition 8 (Appendix A, Terminology).

In our future work, we plan to investigate the performance of our MaxSPRT implementation and compare it with other statistical measures for vaccine safety surveillance. There we plan to incorporate these variations in the performance analysis and describe our findings in detail following (Greene et al, 2011).

### 3.5. Flowchart

Figure 3, shows a flow diagram of the MaxSPRT analysis. It describes the steps from taking input through to signal detection, including possible decision points to get a variation of MaxSPRT utilising the available information and selecting from possible options. Note that our current implementation is focused on the variations of MaxSPRT where baseline information is not known or insufficient. The diagram, however, shows a general flow of processes for most of the possible implementations and variations, including sufficient baseline information known.

**Fig. 3.**
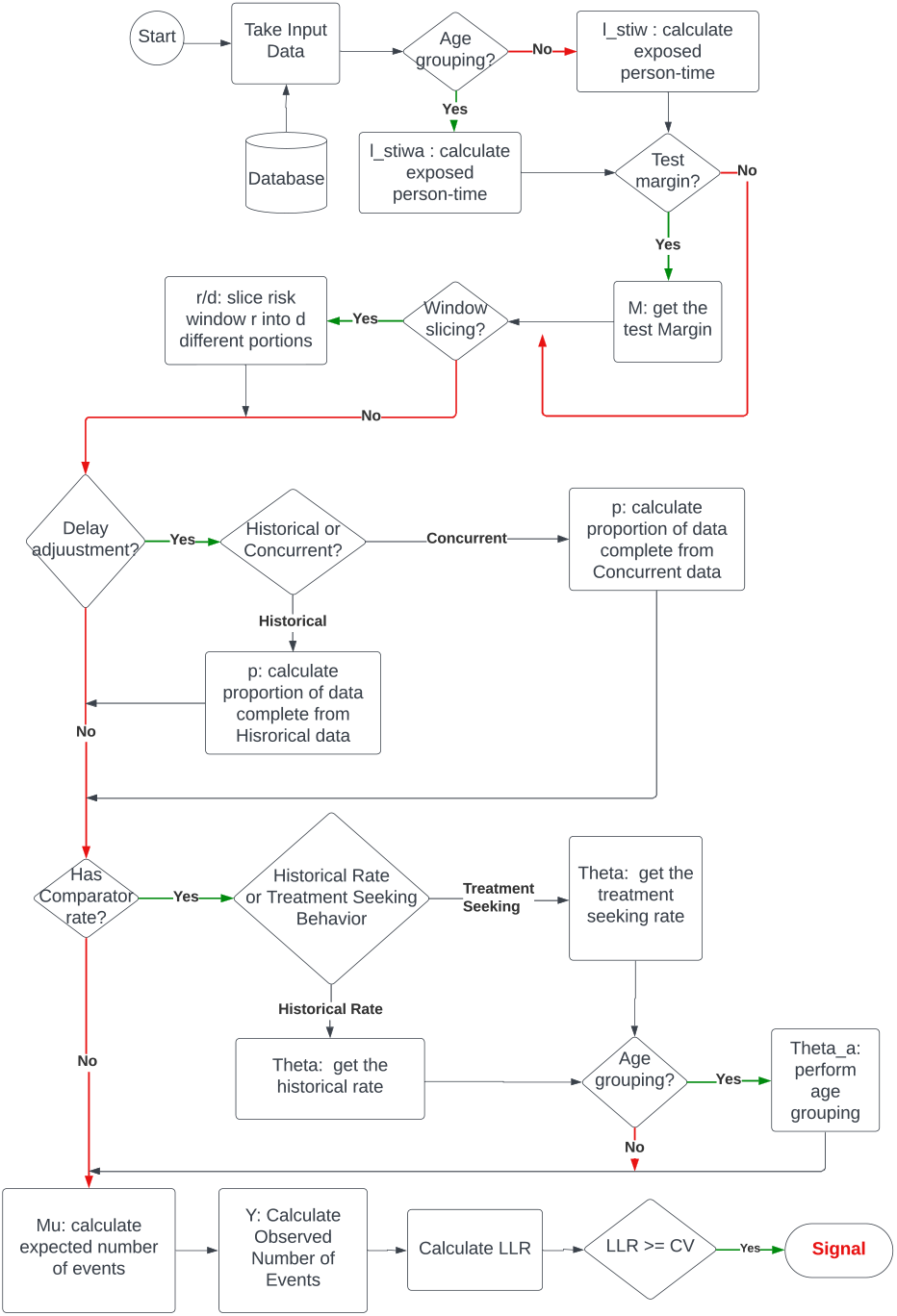
Flowchart of MaxSPRT.

## 4. Discussion

In the previous sections, we introduced the variations of MaxSPRT, the reasons and the factors behind the variations, the challenges associated with particular variations, and how to deal with them. We also explain how we implemented it with maximum flexibility to offer as many variations as possible while minimising editing to underlying code. Our current implementation, however, does not include as yet all possible variations of MaxSPRT, however, our Object-Oriented code can be easily extended with new variations, functionalities, and parameters without breaking any working code or module. This extensibility offers the opportunity to add new variations and devise more effective and adaptable signal detection.

Some additional potential variations that can be added are as follows:

1. Based on different ways of calculating the proportion of data complete, as shown in Table 5, there are five variations using the data available to us. More variations can be added based on the nature, characteristics, source, and purpose of the data.
2. Another prime source of variation in MaxSPRT is obtained through the proportion of data complete calculation based on the data source. Our options are using either concurrent data or historical data. We have added both in our current implementation.
3. Based on the calculation of length of exposure. Currently, we use observed exposed person time. This exposed person-time can be calculated in a different and perhaps a more reasonable way that adds more effectiveness in the calculation of the length of exposure and thus expected number of events.
4. CDC has not used risk window slicing in (Center for Biologics Evaluation and Research, 2021) but Greene et al. used risk window slicing in (Greene et al, 2011). Hence, the risk window slicing can add another variation in MaxSPRT analysis to the conditional MaxSPRT.
5. A new thread to the variations can be weighted average of expected event counts calculated using the formulas used in Definitions 8 (Appendix A, Terminology). This is only possible when baseline data is available. The efficacy of this idea needs extensive attention in terms of theoretical development and experimental analysis.

We also plan to extend our current implementation in various dimensions, such as applicability in surveillance and in other domains, to increase reusability, and enhance capability; towards wider accessibility, performance assessment and comparison, providing soundness in signal detection, and adaptability. We can extend our current implementation and the work on MaxSPRT in the following ways:

### Applicability in surveillance

Integrate MaxSPRT in a Bayesian Network (BN) (Koller and Friedman, 2009; Jensen and Nielsen, 2007) model that is being developed for signal detection within our organisation. Our plan is to build a BN model containing widely used statistical surveillance techniques as integrated nodes. Finally, the BN is expected to perform probabilistic inference (Samiullah et al, 2023; Flores et al, 2004) and find signals more effectively.

### Performance benchmarking

We plan to compare the performance of different variations of MaxSPRT and different statistical surveillance methods and techniques, such as proportional reporting ratio (PRR) (Clothier et al, 2019), Empirical Bayes (Harpaz et al, 2013), CUSUM (Musonda et al, 2008), for safety signal detection. This requires some standards of comparison, and our initial plan is to apply all the variations of MaxSPRT and all other statistical methods under consideration on the same data sample. We can then compare MaxSPRT variations in their capacity to detect signals under particular circumstances. Among the methods that can detect signals, we can rank them based on the time required to detect a signal. We can also check if the generated signal is a false alarm or not, by checking the results among the methods. This develops another way of evaluating the capability of the methods in signal detection. We can then perform the analysis of effectiveness among the methods by calculating precision and recall.

### Providing wider accessibility

Difficulties faced in MaxSPRT implementation can occur due to challenges associated with the range of potential variations; no definite answer to some critical issues; and a relative lack of established literature. Another set of challenges was the verification of the correctness and soundness of the implementation. This motivated us to write this paper and further plan to develop an Application Programming Interface (API) that can be used by researchers to detect signals using their data. Using an open-access API can also assist in verifying implementation by comparing implementation results with the output and scores generated by our API on the same data.

### Adaptability and extensibility

Our current implementation is only for data with Poisson distribution. The next step in our research is to integrate the facility to cope with the data having binomial distribution.

## 5. Conclusions

MaxSPRT is a complex but powerful method for sequential and continuous surveillance. It provides flexibility for implementing in different situations and conducting surveillance with various data limitations. However, its flexibility comes with implementation and operational trade-offs. The associated challenges can make it difficult to implement in some contexts or organisations, which can prevent benefits from being realised.

In this paper, we explained the variations; factors behind the variations; parameters associated with different variations; potential ways of calculating parameters; challenges in calculating parameters; and consequences of using and not using various parameters. We also provided a step-by-step implementation description; demonstrating the corresponding output of each step. The step-by-step discussion also included a detailed description of our data, handling and preprocessing of the data, and our working environments. This paper explained the MaxSPRT surveillance method in a new, different, clear, and intuitive way aiming to support its reproduction by providing mathematical and scientific notations and definitions of the relevant terms that mitigate the gaps between concept and reality. We included a flowchart of the overall implementation method for different variations of MaxSPRT; and also added a hierarchy of the variations to demonstrate the evolution of the methods.

Furthermore, we proposed new measures to assist in the calculation of some parameters. In case there are any special attributes or some missing attributes in our data, we showed in a detailed manner how to deal with the issues and proposed appropriate workarounds as a guideline for reproduction.

We discussed further variations of MaxSPRT that may better capture a signal and our plan to extend the work, aiming to provide greater support to the vaccine safety surveillance community and to the public health surveillance community more broadly. We intend to further the work with the empirical outcomes of the variations of MaxSPRT and other surveillance techniques. Our paper provides a clear road map for future researchers and pharmacovigilance practitioners to implement MaxSPRT not only for vaccine safety surveillance but also for broader adverse drug event sequential analysis and safety monitoring.

## Data Availability

Not applicable

## Declarations

## Acknowledgement

We thank Professor Ann E Nicholson, Faculty of Information Technology, Monash University, Dr. Aishwarya Shetty, Epidemiologist, Murdoch Children’s Research Institute, and Dr. Ben Atkins, Statistical Data Analyst, Murdoch Children’s Research Institute for their support, knowledge sharing, and providing various input from the beginning of the journey through to developing the framework and writing the paper.

## Ethics Approval

This paper does not require ethics approval.

## Consent to Participate

Not applicable.

## Consent for Publication

Not applicable.

## Availability of Data and Material

Not applicable.

## Code Availability

Not applicable.

## Authors’ Contributions

All authors contributed to the study conception. The frst draft of the manuscript was written by Samiullah, and all authors commented on previous versions of the manuscript. All authors read and approved the final manuscript.

### Funding acquisition

Buttery

### Investigation

Samiullah, Clothier, Yin, Munakabayo, Kattan, Mallard

### Methodology

Samiullah

### Project administration

Samiullah, Dimaguila Supervision: Clothier, Dimaguila, Buttery

## Appendix

### A Terminology

#### Definition 1 Study Week

Let *s* be the week at which the MaxSPRT analysis is being performed to find any potential signal for an AEFI on a particular dataset. This week, *s*, is known as study week.

#### Definition 2 Study period

Let *s* be the study week of surveillance through MaxSPRT analysis. The time between the beginning of an observation or surveillance and the study week *s* is known as the study period, i.e., from the 1st week to week *s*, and denoted as [1: *s*].

As an example, if an epidemiologist wants to perform a MaxSPRT signal detection analysis for fever post-influenza vaccination as an AEFI at week 10 of the observation period, then *s*=10. In this case, the study period is Week 1 to 10.

#### Definition 3 Intermediate week

Let *s* be the study week of surveillance through MaxSPRT analysis. Any week, ranging from the start week (time/week of vaccine administration starts) up to the study week *s*, is called an intermediate week and denoted as *t* where 1 ≤ *t* ≤ *s*.

For MaxSPRT analysis, we need to calculate several values in a weekly fashion, from week 1 of the observation week up to the observation/study week *s*. These weeks are known as intermediate weeks and are denoted by *t*. So, *t*=1, 2, …, *s*.

#### Definition 4 Exposed weeks

Let *s* be the study week of a MaxSPRT analysis of a vaccine for an AEFI that has a risk window of length *RW* days. Then, for any vaccinee who was administered a dose of the vaccine on week *t* and having *T T O* days of time-to-onset is in the exposed state for *min*(*T T O, RW*) days starting from week *t* just after the day of the dose administration. The weeks from *t* to *t* + ⌈*min*(*T T O, RW*)/7⌉ are known as exposed weeks for the vaccinee and denoted as *w*.

In other words, exposed weeks of a vaccinee are the weeks after a dose is administered to the person until the reaction/AEFI is observed on them.

As an example, if someone gets a dose on week-2 of the study period, and the risk window is 4 weeks or 28 days, then 2^*nd*^ week of the study period (*t*=2) is considered as the administered week and denoted as *w*=0 at week *t* = 2, week *t* = 3 is the 1st exposed week and denoted as *w*=1, and so on. This continues up to *w*=4 during week *t*=6 of the study period.

#### Definition 5 Time to onset

Let a vaccinee *V* be administered a dose on day *d* (calendar date *D*) of the week *t* and a MaxSPRT analysis being performed in week *s* where *t* ≤ *s*. If the AEFI is observed on that vaccinee in day *d*^′^ (calendar date *D*^′^) of week *t*^′^ where *t* ≤ *t*^′^ ≤ *s*, then the time required for the reaction to start/set is the difference between the two days *D* and *D*^′^ is known as the time to onset and denoted as *T T O* where *T T O* = *interval*(*D*^′^ − *D*).

*T T O* can be expressed in hours or in days in the SAE-FVIC environment/system. As an example, if a vaccinee reported that on the 15th day after the dose the AEFI occurred, then *T T O*=15 days. This is rounded up to a day if *T T O* is given in hours.

#### Definition 6 Risk Window

Let a vaccinee *V* be administered a dose on day *d* of the week *t*. Say, the AEFI is observed on that vaccinee on day *d*^′^ of week *t*^′^ where *t* ≤ *t*^′^ ≤ *s* and *T T O* be the time-to-onset of the AEFI for the vaccinee *V*. The maximum value of (*T T O, RW*) for which the AEFI be considered as an effect of the vaccine is known as the Risk window and is denoted as *RW*.

In other words, the Risk window is the maximum number of days/weeks a vaccinee is at risk of the AEFI after a dose. As examples, some AESIs with relevant risk windows are shown in the following Table 8.

**Table 8.**
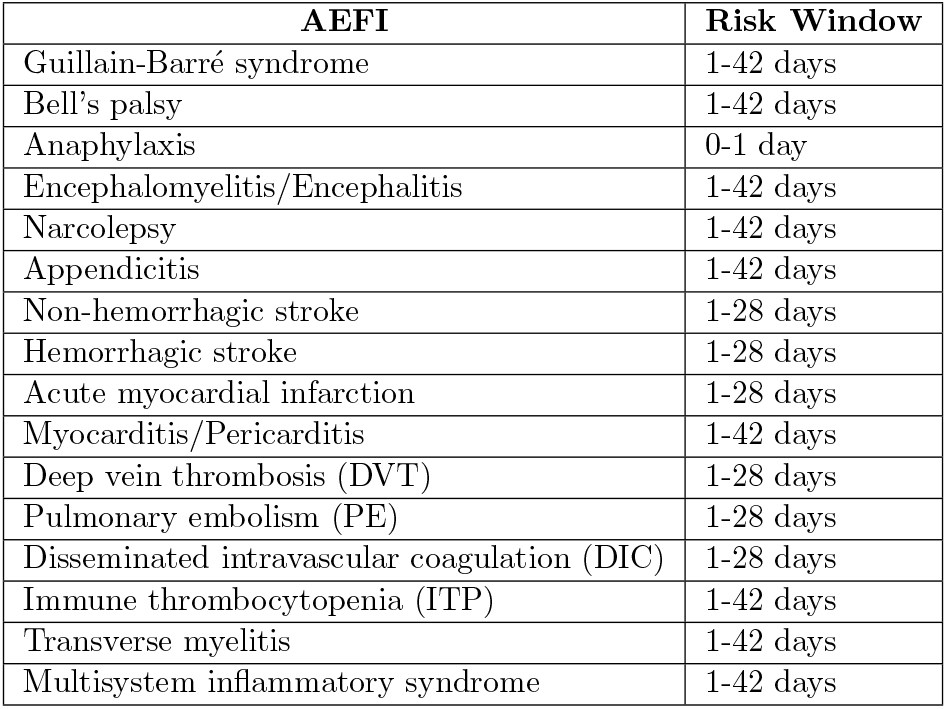
Example of risk windows for potential AESIs (Center for Biologics Evaluation and Research, 2021)

| AEFI | Risk Window |
| --- | --- |
| Guillain-Barré syndrome | 1-42 days |
| Bell's palsy | 1-42 days |
| Anaphylaxis | 0-1 day |
| Encephalomyelitis/Encephalitis | 1-42 days |
| Narcolepsy | 1-42 days |
| Appendicitis | 1-42 days |
| Non-hemorrhagic stroke | 1-28 days |
| Hemorrhagic stroke | 1-28 days |
| Acute myocardial infarction | 1-28 days |
| Myocarditis/Pericarditis | 1-42 days |
| Deep vein thrombosis (DVT) | 1-28 days |
| Pulmonary embolism (PE) | 1-28 days |
| Disseminated intravascular coagulation (DIC) | 1-28 days |
| Immune thrombocytopenia (ITP) | 1-42 days |
| Transverse myelitis | 1-42 days |
| Multisystem inflammatory syndrome | 1-42 days |

#### Definition 7 Observed exposed person-time

Let *s* be the study week of a MaxSPRT analysis and a vaccinee *i* of age group *a* is administered a dose on week *t* and 1 ≤ *t* ≤ *s*. The number of days of a week *w* (*t* ≤ *t* + *w* ≤ *s*) after vaccination and until the reaction/AEFI is set on the vaccinee is known as exposed person-time for the vaccinee *i* in *w*^*th*^ week and denoted as *l*_*stiwa*_.

It is required for calculating the expected number of events, *µ*_*s*_, and represents the length of exposed time for a vaccinee after a dose administration in a person-time unit.

Note, *l*_*stiwa*_ has five terms incorporated in it, namely, *s, t, i, w*, and *a* that corresponds to observation week *s*, dose week *t, i*^*th*^ vaccinee, *w*^*th*^ exposed week and *a* age group in which the vaccinee *i* belongs to. It is defined as the number of exposed days or length of exposure of a person *i* of age group *a* who got a dose on week *t* where *t*^*th*^ week is a week within the study week *s*. Note that for any vaccinee, *w* = 0 means the week the vaccinee *i* was administered a dose and *w* = 1 means 1^*st*^ exposure week of the person.

*l*_*stiwa*_ ranges from 0 to 7. If the dose is administered on the selected last day of a week *t, l*_*stiwa*_ for the week *t* will be 0 and denoted as *l*_*sti*(*w*=0)*a*_ = 0. More specifically, if the person was 7^*th*^ in the list of people reporting for that week of their AEFI and got the dose on the third week of the study period of 10 weeks, then the length of exposure for the first week of the person (or third week of the study for the person) is 0 and denoted as *l*(*s*=10)(*t*=3)(*i*=7)(*w*=0)(*a*=50−59) = 0, where the person is categorised in the age range between 50 and 59. If the risk window of the AEFI is greater than or equal to 7 days, then for the same person the *l*_*stiwa*_ for the next week is *l*(*s*=10)(*t*=4)(*i*=7)(*w*=1)(*a*=50−59) = 7.

Say, for a risk window of 14 days, if the dose was administered on day 2 of a week, then *l*_*sti*(*w*=0)*a*_ = 5, *l*_*sti*(*w*=1)*a*_ = 7 and *l*_*sti*(*w*=2)*a*_ = 2. On the other hand, if the dose was administered on day 4 of a week, then *l*_*sti*(*w*=0)*a*_ = 3, *l*_*sti*(*w*=1)*a*_ = 7 and *lsti*(*w*=2)*a* = 4.

#### Definition 8 Approximate Expected number of events

Following the definition of (Center for Biologics Evaluation and Research, 2021), let *s* be the study week of a MaxSPRT analysis and *t* be any intermediate week and 1 ≤ *t* ≤ *s*. The approximate number of events that are to occur in a week *t* under the null hypothesis, is known as the approximate expected number of events. It is denoted as 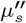 and defined as:

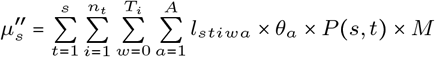

where,

- *s* is a number representing the week when the signal detection analysis is being performed,
- *t* is a number within [1, *s*] denoting any particular week from week 1 up to the study week,
- *n*_*t*_ is the number of vaccinee got a dose on week *t*,
- *i* is any particular individual or vaccinee,
- *T*_*i*_ is the exposed weeks for vaccinee *i* ranging from [1,
- *RW* ],
- *RW* is the risk window length, expressed in the number of weeks,
- *w* is a value in the range [0, *T*_*i*_] where *t* + *w* represents a week under risk where *w* = 0 represents *t* + 0 week of the surveillance which is the dose week of vaccinee *i* and *w* = 1 is the *t* + 1 week in the study and 1st exposed week for the vaccinee,
- *A* is the set of age group strata,
- *a* is the strata of the vaccinee *i*,
- *l*_*stiwa*_ is the observed exposed person-time, and
- *θ*_*a*_ is the per-day comparator rate of age group *a*.
- *M* is the test margin.

Let *RW* be the risk window for the AEFI under consideration for MaxSPRT analysis. If the analysis being performed is in *δ* slices, then all the calculations for MaxSPRT are performed for each slice of the window. For some variations of MaxSPRT, we can split the risk window into several parts. Say, the risk window is 42 days, if *δ* = 6, then for each slice, the number of days = 7.

##### Definition 9 Expected number of events

Let *s* be the study week of a MaxSPRT analysis and *t* be any intermediate week and 1 ≤ *t* ≤ *s*. The number of events that are likely to occur in week *t* under the null hypothesis, is known as the expected number of events and denoted as *µ*_*s,t*_.

It is one of the two parameters required to calculate *LLR*. Calculating *µ*_*s,t*_ is tricky and complicated. Estimating this value requires approximation in person-time. There are different approaches that can be adopted based on available data and information to estimate the value of *µ*_*s,t*_.

##### Definition 10 Observed number of events

The number of events that occurred under the risk window in week *t* and reported as well as observed by week *s*, is known as the observed number of events and denoted as *Y*_*s,t*_.

This is another parameter for calculating *LLR*. It counts the weekly events within the risk window and is reported and observed by the study period.

##### Definition 11 Historical comparator rate

Let a MaxSPRT analysis be performed for an AEFI due to a vaccine. Say, in previous *y* years, *n* people were observed to be affected by the AEFI out of *N* people, then the historical comparator rate is defined as the ratio of the observed number of affected people by the AEFI, i.e., 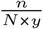 and denoted as *θ*.

It is the value in person-time unit required to calculate the expected number of events, *µ*_*s*_, representing the number of people per year that suffer from an AEFI. It is calculated retrospectively from the previous years’ data.

##### Definition 12 Per-day comparator rate

Let *θ* be the historical comparator rate in previous *y* years obtained by observing *n* people to be affected by the AEFI out of *N* people, then the per-day comparator rate is defined as the ratio of the observed number of affected people by the AEFI in a day, i.e., 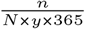 and denoted as *θ*^′^.

In the case of age grouping stratified MaxSPRT analysis, the historical comparator rate and per-day comparator incidence rate may also vary. In order to address that, these rates for any age group *a* are denoted as *θ*_*a*_ and *θ*^′^, respectively. There are several ways of calculating the rate where two of the plausible ways are:

- the rate of the AEFI within the general population
- healthcare-seeking behaviour for AEFI following vaccination

##### Definition 13 Observation delay

Let *s* be the study week when a MaxSPRT analysis is being performed, *V* = {*v*_1_, *v*_2_, …, *v*_*n*_} be the set of vaccinees reporting the AEFI, *T* = {*t*_1_, *t*_2_, …, *t*_*n*_} and *T* ^′^ = {*t*_1_ ^′^, *t*^′^_2_, …, *t*_*n*_^′^ } be the set of corresponding AEFI observation weeks and AEFI reporting weeks of the vaccinees in *V*, respectively, where 1 ≤ *t*_*i*_ ≤ *t*^′^_*i*_ ≤ *s*, 1 ≤ *i* ≤ *n*. Then the time elapsed between the dose administration and AEFI reporting is known as observation delay and denoted as, *δ* = *interval*(*T* ^′^, *T*).

AESIs may not be promptly reported in real time due to various factors. Even when reported, there can be delays in recording AEFI cases, a phenomenon referred to as observation delay. These delays result in a reduced amount of observed data compared to the actual amount during analysis, potentially leading to an underestimation of expected event numbers and a risk of missing signals. To mitigate this issue, cMaxSPRT incorporates an approximation technique involving the calculation of the proportion of complete data (the ratio of observed to expected data). This concept is defined in the following Definition 14.

##### Definition 14 Proportion of data complete

Let *s* be the study week when a MaxSPRT analysis is being performed and *δ* be the observation delay in the reports. Then, the proportion of data complete at week *s* is the ratio of reports of week 1 ≤ *t* ≤ *s* that are available by week *s* due to observational delay and total expected reports for week *t*. It is denoted as *p*(*s, t*).

As an example, at week *s*=10, we observe 6 reports of week *t* = 5, and it is expected that 10 reports to be noted for week *t* = 5. That means, only 60% of week *t*=5 data or report is available. This is denoted as *p*(10, 5) = 0.6.

There are several ways or heuristics to calculate this value. A retrospective analysis of historical data can be an option to compute this metric.

At the time of analysis, we may not obtain all the reports in time due to various reasons. Addressing this factor while estimating the expected number of events is crucial. Otherwise, we may overestimate the expected number of events. To address this issue and adjust the expected number of events, CDC (Center for Biologics Evaluation and Research, 2021) proposed the following equation.

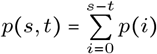

We explained the calculation techniques and challenges of this metric in the paper in Section 3.2.3.

##### Definition 15 Cumulative expected number of events

Let *s* be the study week when a MaxSPRT analysis is being performed and *µ*_*s,t*_ be the expected number of events (reports) to occur in weeks 1 ≤ *t* ≤ *s*. Then the cumulative expected number of events is denoted by *µ*_*s*_ and defined as:

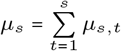

##### Definition 16 Cumulative observed number of events

Let *s* be the study week when a MaxSPRT analysis is being performed and *Y*_*s,t*_ be the observed number of events (reports) occurring in weeks 1 ≤ *t* ≤ *s*. Then the cumulative observed number of events is denoted by *Y*_*s*_ and defined as:

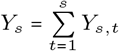

It represents the number of AESIs occurring within the exposed risk window. It can be easily calculated by counting the cumulative sum of AEFI counts occurring weekly straightway. In order to find the number of observed events up to week *s*, we have to simply accumulate the number of events occurring weekly.

##### Definition 17 Log-likelihood ratio

Let *s* be the study week of a MaxSPRT analysis where *t* be any intermediate week and hence 1 ≤ *t* ≤ *s*. Also, let *µ*_*t*_ and *Y*_*t*_ be the cumulative approximate expected number of events and cumulative observed number of events in the study, respectively, up to week *t*. Now, according to (Kulldorff et al, 2011), the Loglikelihood ratio of week *t* be denoted as *LLR*_*t*_ and defined as follows:

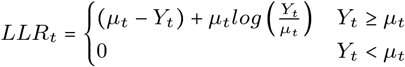

##### Definition 18 Critical value

Let *s* be the study week of a MaxSPRT analysis and *LLR*_*t*_ be the Log-likelihood ratio for any intermediate week 1 ≤ *t* ≤ *s*. According to (Kulldorff et al, 2011), the value of *LLR*_*t*_ (or *µ*_*t*_ if the analyser chooses this over *LLR*_*t*_) that makes the alternative hypothesis true is defined as a critical value and denoted as *CV*.

There are two ways of detecting a signal. Firstly, if the cumulative number of events observed at any time is beyond a prespecified number, then there is no need to perform any MaxSPRT analysis and the null hypothesis can be rejected straightaway. Otherwise, at any week *t*, if the *LLR*_*t*_ (or *µ*_*t*_) is beyond the corresponding critical value *CV* expressed in likelihood (or, expressed in event count), then there is a signal, that is, 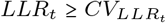, where 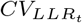 or 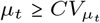, where 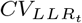 or 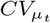 is the critical value for *µ*_*t*_ being the cumulative expected number of events for week *t*.

Kulldorff et. al. (Kulldorff et al, 2011) proposed the technique, calculation, and uses of *CV* s for signal detection. They also developed an R sequential package with all functionality native to MaxSPRT calculation. If someone uses R, they do not need to reinvent the wheel for computing CV and other parameters required for MaxSPRT analysis. Moreover, in their paper they have given *CV* s for up to 1000 expected number of events, for the reason that if someone wants to use a *CV* for signal detection, they do not need to compute *CV* s again.

In this paper, we offer a detailed explanation of the steps and challenges associated with the implementation of MaxSPRT on SAEFVIC’s reporting database. SAEFVIC is the jurisdictional vaccine safety surveillance service of Victoria, Australia. Its AEFI and AESI reporting database (Clothier et al, 2011) does not currently possess dose administration information. Dose administration data is collected from the Australian Immunisation Register. At the time of writing, SAEFVIC only had access to COVID-19 vaccines, in near real-time. Hence, the denominator of the MaxSPRT calculation - the expected number of events - requires estimation or conditioning to deal with its associated uncertainty. Therefore, in our implementation, we use the concept of cMaxSPRT, similar to the approach adopted by the US CDC (Center for Biologics Evaluation and Research, 2021) for surveillance of COVID-19 vaccine safety using insurance claim data. In both cases, the denominator is not known in real-time and at least not during the analysis. Being able to use a statistical method e.g. MaxSPRT in the absence of dose distributed denominator is therefore a practical advantage for spontaneous reporting systems to function as stand-alone signal detection systems.

### B Data Processing and Environment Setup Implementation

In this section, we have explained the steps of implementation in a particular surveillance setting (in our case SAEFVIC).

#### B.1. Preprocessing

SAEFVIC data are prepared prior to processing. The preprocessing involves de-identification, noise handling, and missing data management. We have used the features of the MaxSPRT framework to deal with the pre-preprocessing challenges, especially, the missing data management. We allow analysers to opt-in methods to fill in missing values with some appropriate alternative values or simply discard the rows/records with missing values.

To justify filling in missing values, which is questionable from the point of view of epidemiology, this is logical from a data analysis perspective in certain scenarios. As an example, if we discard the records for some missing values in gender but the gender-specific stratification is not required in the calculation or analysis, then removing that record is not worthwhile. Rather, discarding the value adds bias to the numerator and denominator calculations. However, it is up to the user of the framework to decide whether to use the filling-in method or not.

We first de-identify the data to preserve the privacy of the reporters and vaccinees. In the framework, we allow for the de-identification of the data, which is quite straightforward, by removing all demographics and identifiable information about the vaccine and the reporter. The analyser can also mention if any further information needs to be removed for the de-identification other than the demographics. In the deidentification process, we assign a unique identifier to each report for record-keeping purposes. We keep the columns as specified in Table 2.

Next, we dealt with noisy data. We treat noisy data in a special manner by initially considering them as missing values. In this paper, noisy data are defined as values or inputs susceptible to error due to human and system error. First, we run a routine data cleaning for logic consistency, missing values, and filtering process, where any value that is beyond the range and domain of the corresponding data attribute is identified as noisy data. We first replaced a noisy value with missing values. Then treated them the same way as missing values.

In the third preprocessing step, we dealt with missing values. A common practice of dealing with missing values in data processing is to delete rows with noisy or missing values. However, in the case of vaccine safety surveillance, every report is important, so it is necessary to manage, instead of removing, missing records. We needed to carefully deal with missing records and use effective ways to fill them in. Of course, fill-in methods sometimes affect some important properties of the data such as mean, median, mode, standard deviation, and variance of the data. Therefore, the choice of the right fill-in method that has the least negative impact on the data is essential. Nevertheless, we leave it to the analyser to use fill-in, discard the record, or use any specific way of dealing with the missing and noisy data. In case, the analyser chooses to fill in missing values, then one of the following could be used: (i) default value/s, (ii) data-type specific alternative value, (iii) the analyser provided alternative values, or (iv) a method to get appropriate alternative values provided by the analyser.

To the best of our knowledge, there is no best or single approach to dealing with missing values. Hence, in our framework, we allowed several methods to fill in missing values. An end user of the framework can opt into a specific method through parameterisation if they want to override the default one in their MaxSPRT analysis. Moreover, our data processing unit is capable of accepting new methods of filling in the missing values. The methods integrated into the current implementation that are used to fill in missing values in our data are actually the available methods in the Python Pandas package for fill-in such as “replace” with mean, median, mode, or some specific value like zero; and “interpolate” which also comes with a handful of options^1^.

To explain the preprocessing process on our data, the following steps are taken. As an example, for different columns, we have used different default fill-in methods for illustration purposes. For missing “Age” and “TTO” values, we used linear interpolation as default. For “Sex” attribute, the missing values can be either replaced by randomly chosen value between “M” and “F” or as per the AESI we can decide if bias for a particular “Sex” is to be considered. By bias, we mean, some AESIs are more common or only for one sex (male or female) in comparison to the other. For treating missing values of “Time to vaccine”, “Reporting time” and “Submission time”, we converted the times to double precision real numbers and then applied linear interpolation. However, this part is quite tricky and crucial, as these values will be used extensively in calculating most of the important terms in MaxSPRT. Also, there is an inherent order of the three values by definition, and the order is *T ime to vaccine* ⪯ *Reporting time* ⪯ *Submission time*. Conducting the fill-in should not violate this order constraint. There is another constraint on “Time to vaccine”, “Reporting time”, and “TTO” that is *Reporting time* − *T ime to vaccine* ≤ *T T O*. Therefore, the interpolation of “Time to vaccine”, “Reporting time”, and “TTO” should satisfy this constraint as well.

We transformed the three timestamp values while dealing with missing values. In addition, we ran another iteration of data transformation on our data before we started processing them for analysis. The transformation includes a common and unique format of timestamps for all three timestamps we have used in our data, and *T T O* values are rounded up to the nearest integer value.

To perform age-based and sex-based MaxSPRT analysis, we use age and sex columns. However, age-based analysis requires binning (or grouping or stratification of ages). We considered stratification as a preprocessing step. Here, we added a column to the data, “Age-Strata” having nine different strata, namely 0–9, 10–19, 20–29, 30–39, 40–49, 50–59, 60–69, 70–79, 80+. This grouping can be done on more granular levels and also in a dynamic way rather than the static grouping as shown here. The alternative ways of age grouping also added variations to the analysis, as we mentioned before. Hence, we kept the scope to feed in new binning techniques as well as new or different age strata in our system.

Finally, we discarded the reports where *T T O* is beyond the risk window, and this is an important step. Note that this is applicable if it is decided that the reactions that were observed after the specified and well-defined risk window are not considered in surveillance.

As a pre-processing step of the data, we assessed that the values/rates need to be scaled by the length of risk window. This is because these values are computed in terms of persontime on a scale of 100,000 people per annum, and the risk windows are expressed in days. As an example, if the rate is 5.0 for 100,000 people-year and the risk window is 40 days long, scaling it down to 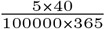 or 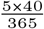 for 100k people is required to be used in MaxSPRT analysis.

Once the scaling is performed, we call them a per-day comparator to estimate the expected number of events. This is referred to as the expected rate of AEFI approximated for the currently vaccinated population which is not known, at least during the time of analysis.

We performed age-based stratification of the whole MaxSPRT analysis. Hence, this parameter offers another layer of computation where the rates vary for different age groups. In the case of no age-based stratification, we assumed a single and common rate for all age groups. This allows for facilitating the use of age groups and avoiding the age groups without any major reworking in the implementation, especially in the other modules of the implementation that are developed to allow age grouping in mind. Note that in cases of age grouping, all the calculations are performed at a more granular level and all the parameters used, including this one, are taken/calculated based on the age ranges of the groups.

Interestingly, in the case of some parameters that are not available for age groups, we used the common and general values for all ages. In our current implementation, we have allowed both, adding age-based and general values for this parameter. However, for brevity and simplicity, this paper uses a single and general value for the parameter.

#### B.2. Processing of SAEFVIC Data

After the preprocessing of data, we started calculating the metrics that are required to perform MaxSPRT analysis and detect signals. The analysis is required to be performed in a well-defined frame of weeks for the sake of tracking the computation and facilitating weekly analysis. As a next step, we need a set of continuous windows in the analysis, where the data is divided into different windows of weeks based on their occurrence dates. Our implementation does not rely on any explicit length of the analysis or size of the data frame, or count of windows in the data frame. Moreover, our current implementation allows the analysers or epidemiologists to set their preferred start date, end date, length of analysis or count of windows in the frame.

In this task, the main challenge was to take the starting date of the first week. Then the formation of the windows was quite straightforward. The first window was formed by adding 7 days to the start date of the analysis. This gives us another date. Hence, our windows are defined by the start and end date of the week (does not matter what day of the week it is). Then, adding 1 to the last date of the first week gives us the first date of the second week. We make a set of windows each of 7 days or 1 week long by continuously following the steps. A point worth noting is that the start date of analysis is by default taken from the data by finding the earliest vaccination date. This can also be replaced by the earliest report date or report submission date. On the other hand, the frame ends with a week having an end date that is not later than six days from the latest date of reporting found in the data. Hence, our implementation does not rely on any explicit length of the analysis, the size of the data frame, or the count of windows in the data frame. Moreover, our current implementation allows analysts or epidemiologists to set their preferred start date, end date, length of analysis, or count of windows in the frame.

Finally, while computing various matrices like *p*(*s, t*), *l*_*stiwa*_, *Y, µ*, and *LLR*, the data is further processed and new columns are added to it as required. We described this further processing and amendment of the columns to the data later, in Appendix B.

#### B.3. Environment Setup and tools

Kulldorff et al (Center for Biologics Evaluation and Research, 2021) and Greene et al (Greene et al, 2011) recommended the use of R programming language as there is a package called “Sequential” in *R* developed especially for sequential analysis and contains all the required functionalities for MaxSPRT and other similar continuous and sequential analyses. However, our review of the literature and domain reveals that the main dependency of MaxSPRT implementation on R is to calculate critical values, *CV* for corresponding *µ* with a proper *α* spending plan (Silva et al, 2019). This *α* spending plan will tune the threshold of *α* value automatically based on the situation to detect a signal as soon as possible with as minimum type-1 error as possible. However, Kulldorff et al describes in (Kulldorff et al, 2011) that if someone wants to use the table of *CV* s for a specific *α* (note, the table contains *CV* for three different values of *α*, namely, 0.01, 0.05, and 0.1) and for *µ* of up to 1000, they do not need to regenerate the *CV* s.

We have used Python and various packages (e.g., Panda) of Python that facilitate easy data handling and extensibility. For better error handling, extensibility, and code reuse; and to follow software engineering best practice and principles, we adopted the Object-Oriented Programming style. This allows new methods to be injected easily into the implementation. As an example, “R” codes can be easily integrated by embedding into the Python code segment. Hence, the “R” sequential package and other required “R” codes to deal with *CV* calculation and *α* spending plan can be added as an embedded module in the python implementation of MaxSPRT. We have integrated the “R”-scripting facility and embedded the “Sequential” package in our current implementation to provide maximum facility to the analysers.

Our choice of Object-oriented programming style also helps in implementing the different variations of MaxSPRT by adding more than one method for the same measure/metric like *P* (*s, t*), *θ*, and *l*_*stiwa*_. Therefore, in the later stage especially during MaxSPRT analysis different variations of MaxSPRT can be designed by just choosing parameter values and more variations can be added without breaking any working and existing code/module.

### C Demonstration of *l*_*stiwa*_ calculation

In Table 9, we provide a visual demonstration of the calculation of *l*_*stiwa*_ and how it is calculated for each person. As an example, vaccinee *V*_1_ got a dose on the 1^*st*^ day of week-1. Their *T T O* was 5 days. Hence, the person was exposed from day 2 to day 6 of week-1. Hence, *l*_*stiwa*_ of the person is denoted as *l*_*sti*(*w*=0)*a*_ and calculated as 5. Also, vaccinee *V*_5_ got a dose on the 5th day of week-1. Their *T T O* was 11 days. Hence, the person was exposed from day 6 of week-1 to day 2 of week-3. Hence, the *l*_*stiwa*_ of the person for weeks 1, 2, and 3 are denoted and calculated as *l*_*sti*(*w*=0)*a*_ = 2, *l*_*sti*(*w*=1)*a*_ = 7 and *l*_*sti*(*w*=2)*a*_ = 2, respectively. At the end, the last row of the table shows the weekly *l*_*stiwa*_.

**Table 9.**
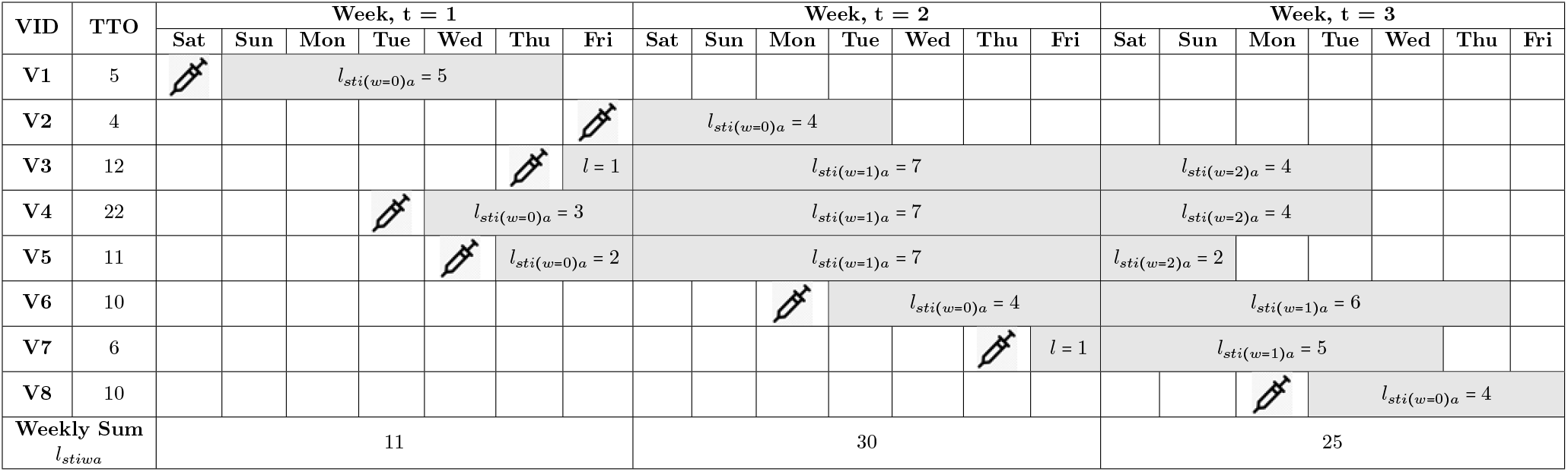
Demonstration of *l*_*stiwa*_ calculation and exposed person-time

Similar to the observed event count, we have added additional columns for *l*_*stiwa*_ for each age group *a*, and for weekly *l*_*stiwa*_ by adding all age-group based *l*_*stiwa*_ for a particular week *t*. As an illustration, Table 4 shows the computed weekly *l*_*stiwa*_ and age-strata based administration of *l*_*stiwa*_.

### D Variations in *p*(*s, t*) calculation

To calculate the ratio for calculating *p*(*s, t*), we need to know the total number of reports for the week. In the literature, this ratio is obtained from historical data. On the other hand, it is quite challenging to get the ratio in case the denominator is not known. Estimating the ratio without a proper or exact denominator needs an approximation technique.

One of the approximation techniques would be to take the total reports found so far as a denominator to compute the ratio. However, this technique would not be sufficient to approximate the ratio properly. Because, as shown in Table 5, it is quite possible that we have more information about earlier weeks and less information regarding later weeks. However, the ratio indicates a wrong proportion of data complete, that is, more for later weeks and less for earlier weeks, unlike a real scenario. In such cases, earlier week ratios will be more accurate than the later weeks. We can estimate the ratios of later weeks from the earlier weeks to deal with this challenge.

Our idea is to estimate a certain number of later weeks from the remaining weeks’ *p*(*s, t*). As an example, we can compute the last 50% of weeks’ *p*(*s, t*) from all previous weeks’ *p*(*s, t*). There is scope to take lesser or greater than 50%. We propose the following equation to calculate the proportion with a certain number of weeks’ *p*(*s, t*) to be estimated.

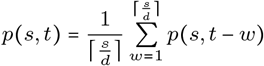

Here we estimate the *p*(*s, t*) for week *t* by taking the average of the *p*(*s, t*^′^) where *t*^′^ is the first half of the study weeks taking the division or partition number, *d* = 2. We can substitute this equation with some other more appropriate estimator, as well as we can tune the equation by changing the values of *d*. As an example, if *d* = 3, the equation will take the average of the 1st one-third weeks’ *p*(*s, t*).

Table 10 shows the possible ways of calculating *p*(*s, t*) for the delay distribution of the reports shown in Table 5. All five different approaches shown in the table for computing *p*(*s, t*) do not comply with CDC’s conclusions that the earlier weeks will have more data complete than the later weeks, but they also have different logic and merits. To choose the best and most effective one, further empirical assessment is required.

**Table 10.** Demonstration of Different *p*(*s, t*) Calculations

| For s = 10 |  |  |  |  | Using Historical Data |  |  |  | Using current data |  |  |  |
| --- | --- | --- | --- | --- | --- | --- | --- | --- | --- | --- | --- | --- |
| t | Expected Count (ER) | Accumulated Count (AER) | Observed count (OR)<br>[up to S] | Accumulated count (AOR) | Idea 1 |  | Idea 2 | Idea 3 | Idea 4 |  | Idea 5 |  |
| | | | | | p(i) | p(s, t) | p(s, t) | p(s, t) | p(i) | p(s, t) | Hazel's $p(s, t)$ | |
|  |  |  |  |  | [proportion of data arriving at week i] | [sum of all p(i) from current to last week] | [AOR / AER] | [AER / Total ER] | [proportion of data arriving at week i] | [sum of all p(i) from current to last week] | expectation: OR/ER (1.B) |  |
| 3 | 6 | 6 | 6 | 6 | 6/332 = 0.0181 | 1 | 6/6 = 1 | 6/332 = 0.02 | 6/265 = 0.02 | 1.00 | 6/6 = 1 |  |
| 4 | 25 | 31 | 24 | 30 | 25/332 = 0.075 | 0.98 | 30/31 = 0.97 | 31/332 = 0.09 | 24/265 = 0.09 | 0.98 | 24/25 = 0.96 |  |
| 5 | 68 | 99 | 58 | 88 | 68/332 = 0.205 | 0.91 | 88/99 = 0.89 | 99/332 = 0.30 | 58/265 = 0.22 | 0.89 | 58/68 = 0.85 |  |
| 6 | 72 | 171 | 62 | 150 | 72/332 = 0.217 | 0.70 | 150/171 = 0.88 | 171/332 = 0.52 | 62/265 = 0.23 | 0.67 | 62/72 = 0.86 |  |
| 7 | 88 | 259 | 78 | 228 | 88/332 = 0.265 | 0.49 | 228/259 = 0.88 | 259/332 = 0.78 | 78/265 = 0.29 | 0.43 | 78/88 = 0.89 |  |
| 8 | 21 | 280 | 19 | 247 | 21/332 = 0.063 | 0.22 | 247/280 = 0.88 | 280/332 = 0.84 | 19/265 = 0.07 | 0.14 | 19/21 = 0.91 |  |
| 9 | 25 | 305 | 9 | 256 | 25/332 = 0.075 | 0.16 | 256/305 = 0.84 | 305/332 = 0.92 | 9/265 = 0.03 | 0.07 | 9/25 = 0.36 |  |
| 10 | 27 | 332 | 9 | 265 | 27/332 = 0.081 | 0.08 | 265/332 = 0.8 | 332/332 = 1.00 | 9/265 = 0.03 | 0.03 | 9/27 = 0.33 |  |
| Total | 332 |  | 265 |  |  |  |  |  |  |  |  |  |

### E Parameters for MaxSPRT analysis

This section introduces some parameters required to perform MaxSPRT analysis for continuous surveillance using Sequential package (Silva and Kulldorff, 2024). Most of these parameters are explained in the manual (Silva and Kulldorff, 2024) with example scenarios and values. These parameters play a very important role in the analysis.

#### Sample size

A non-zero upper limit of the expected number of events under the null hypothesis is referred to as Sample size. This value is needed to early terminate the analysis when the observed number of events outnumbers this value under the null hypothesis (i.e., the null hypothesis is not rejected yet). There are various available methods in the package to estimate an optimised sample size that reduces the estimated time to signal as well as with different values of type-I error, relative risk, and statistical significance.

#### Statistical significance level (*α*) and alpha spending plan

Statistical significance assesses whether the observed risk is not occurring due to random variation. The significance level *α* focuses on controlling Type-I errors and influences statistical significance in the analysis. MaxSPRT allows the defining or controlling of the alpha spending plan by the analyser. There are parameterisable methods to conduct an analysis using MaxSPRT where a specific plan to spend *α*, such as, uniform in every analysis period, or conservative or generous at the beginning and changing exponentially or logarithmically in spending *α*) can be executed. As per the plan, a certain amount of alpha will be spent in each analysis period (where period means weeks if the data is stratified into weeks and analysed weekly). *ρ* is a positive number used for the power type alpha spending function.

#### Statistical Power

Statistical power measures the probability of identifying the risk if it is truly present. It focuses on controlling Type-II errors and is influenced by sample size and significance level.

#### Relative risk

The relative risk *RR* is the ratio of incidence rate in exposed and unexposed populations. In other words, this is the ratio of observed and expected rates of events. It is required that *RR* > 1, because, with *RR* = 1 means there is no difference between observed and expected incidences and hence no risk due to the vaccine. The higher the value of *RR* the more the risk is and the smaller will be the sample size if statistical significance and alpha remain constant.

#### Minimum number of events

In vaccine safety surveillance, normally a minimum number of events need to be observed before rejecting the null hypothesis and marking the vaccine unsafe. This minimum number of events must be positive and should be 4 (Kulldorff and Silva, 2017).

## Footnotes

1 Interested readers are recommended to see Pandas manual or https://www.geeksforgeeks.org/working-with-missing-data-in-pandas/.

